# Factors to Perceived Quality of Community Based Health Insurance Utilization Among Admitted Patients at St. Paul’s Hospital Millennium Medical College and AaBET Hospital, Addis Ababa, Ethiopia

**DOI:** 10.1101/2024.10.16.24315615

**Authors:** Dessalegn Keney Guddu, Yared Lasebew Asres, Abrham Getachew Dullo, Tefaye Getachew Shawel

## Abstract

**Background:** Community based health insurance(CBHI) was introduced by the Ethiopian government in 2011 with the aim to improve access and quality of health care to the poor informal workers. But studies show that CBHI scheme based service quality is also reported to be low.On the other hand, there has been scarcity of data on barriers to quality of service utilized particularly at hospital level by the insured patients.

**Objective:** The aim of this study was to identify the barriers to good quality of community based health insurance utilization among admitted patients at St. Paul’s Hospital Millenium Medical College and AaBET Hospital, Ethiopia.

**Methods:** Cross sectional mixed design implementing quantitative from chart review as well as qualitative study using phenomenological design using focused group discussion and indepth interview involving the concerned CBHI stake holders was conducted from June1-July15,2023. Opendata kit (ODK) was used for quantitatie data collection and SPSS V25 and NVIVO V12 were used for data analysis.

**Results:** Total of 396 clients participated in the study. The mean age of study particiapnts was 43.64± 14.3 years. over all, 72.7% of admitted patients have got good quality community-based health insurance services.Factors significantly affecting quality of CBHI utilization were a clean and attractive hospital environment [AOR = 2.77:95% CI (1.24–6.165)], satisfied with community based health insurance [AOR = 2.45:95% CI (1.11–5.39)] at enrollment, good knowledge of the CBHI scheme [AOR = 1.97:95% CI (1.2– 3.23)], adequate availability of information on CBHI services in hospital [AOR = 2.37:95% CI (1.34–4.21)], higher family monthly income between 5251 and 7800 [AOR = 1.97:95% CI (1.2–3.23)], income more than 7801 birr [AOR = 5.3:95% CI (2.32–10.23)] respectively. The barriers to good quality utilization identified with qualitative exploration include difficulty in accessing hospital service areas(inconvenient) and information, limitation on the type of service provided by the scheme, poor knowledge of patients, overcrowding, long waiting times for chart activation, shortage of drugs and inpatient beds

**Conclusion and recommendation:** This study determined that the magnitude of good quality community-based health insurance services utilization was moderate compared to previous studies.Policymakers and stakeholders should improve monitoring of quality of CBHI services and further multicenter studies are necessary to improve service quality in public hospitals of Addis Ababa.

## 1. Introduction

Health insurance is a formal pre-paid financial mechanism that allows cross-subsidization of the poor by the rich and the sick by the healthy to risk pooling and sharing resources among different willing members. Community based health insurance(CBHI) scheme covers the poor and informal workers while social health insurance is applicable to formal employee (1)

The World Health Organization has endorsed a community-based health insurance scheme, as a shared financing plan to improve access and quality to health services and ensure universal health ccoverage(UHC) especially a mong the poor informal workers(2). Asystemic review in 31 subsaharan Africa including Ethiopia conducted on 111 studies (2000-2020) has reported pooled annual incidence of catastrophic health expenditure of 16.5% which creates vicious cycle of poverty,poor access and poor quality of health care (3). Government of Ethiopia launched CBHI scheme in 2011 in 4 regions which was latter expanded through out the country (4,5) with a vision of reaching 80% of districts and 80% of its population by 2020(6).How ever, poor quality of health services to the insured is threatening sustainability of the CBHI scheme since it can discouraged further enrollment and enhance dropout rates (7).

The mere presence of CBHI scheme system or even enrollement to the scheme doesn’t justify the optimal health care utilization because of challenges during delivery health of care at health facilities. Infact, few studies have shown worse quality of care among insured as compared to uninsured. According to a systemic review conducted in Africa and Asia,weak evidence points to a positive effect of both SHI and CBHI on quality of care and social inclusion(8)

St. Paul’s Hospital Millenium Medical College (SPHMMC) is providing health care for CBHI eligible patients since 2018 and the number of patients using it per month has gradually increased and reached an average of 10000 by 2022 (taken from SPHMMC CBHI registry). AaBET Hospital which is an affilation of SPHMMC is also providing the CBHI scheme based service since 2019 reaching 1500 beneficiaries per month by 2022.Despite presence of low level of utilization of CBHI and compromised quality of CBHI scheme based services in few studies (2,9), there is paucity of evidences on barriers to quality of health services especially at hospital level in Ethiopia.Therefore, this study aimed to identify the level of quality of CBHI scheme based services and its barriers at SPHMMC and AaBET Hospital.

The conceptual frame work was adapted from review of literatures on challenges againist quality of CBHI utilization by clients at health facility level. The challenges were classified to those related to client characterstics,hospital system of health care delivery and health care workers behavior while serving CBHI insured clients.

**figure I.**
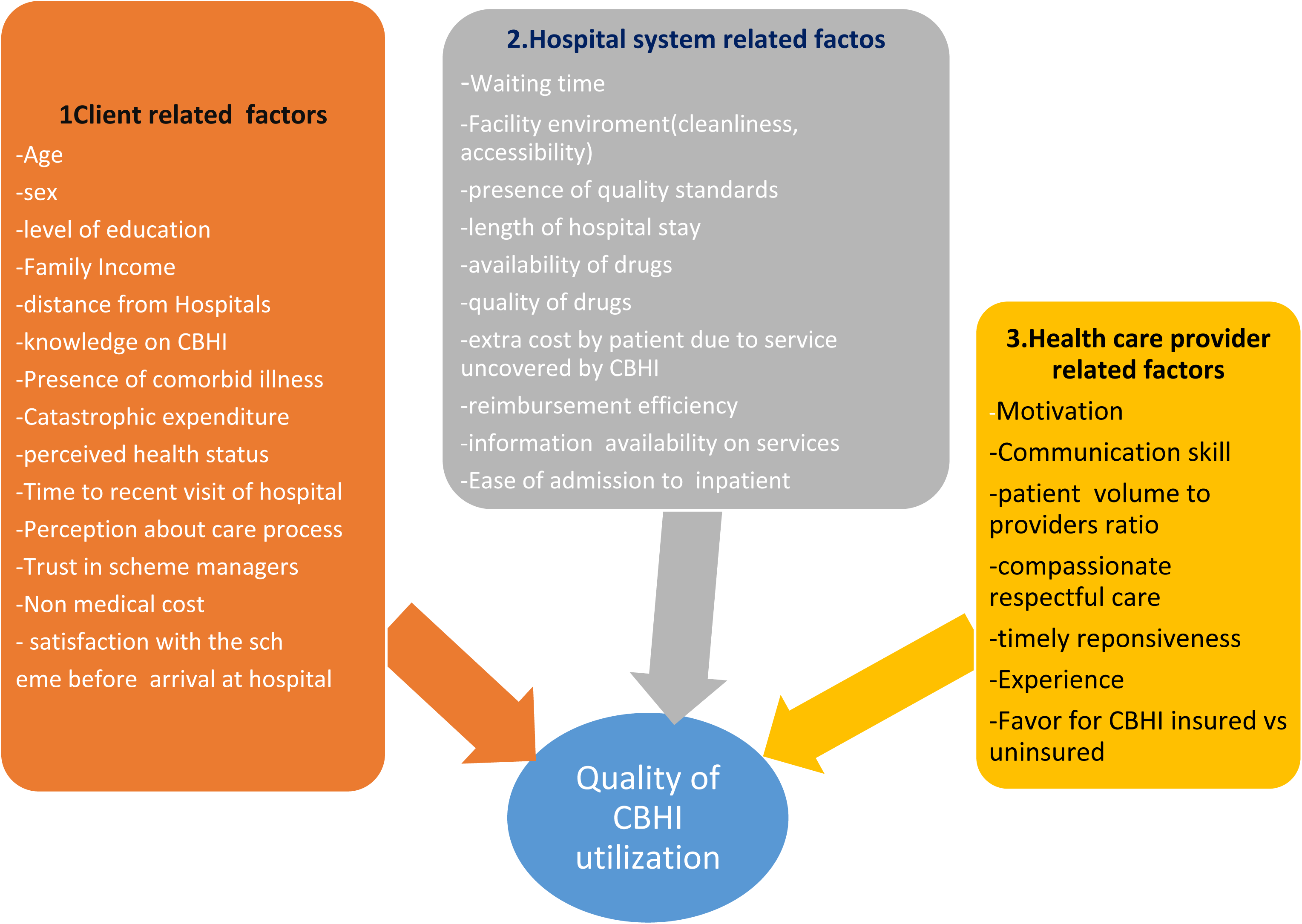
Conceptual fra,ne work of dependent and independent variables.

## 2. Methods and materials

### 2.1 Study Setting /area and period

This study was conducted at St Paul’s Hospital Millennium Medical College(SPHMMC) and AaBET Hospital inpatient departments in Addis Ababa, Ethiopia.. SPHMMC is a specialty and subspecialty service center with and a catchment population of more than 5 million, annual average out patients service of 500,000, 751 bed capacity, 10,000 maternal deliveries and adult emergency flow of more than 33,000.

The hospital is providing CBHI based health service for the insured clients since 2018 with more than 10,000 patients /month (SPHMMC CBHI registry, Dec 2022). How ever, this study was conducted on admitted insured clients which are around 1100 per month in average in 2022.

AaBET Hospital is abranch and an affiliate of SPHMMC which is located at distance of 2.5 km from SPHMMC and running 35 emergency beds and 147 inpatient beds. AaBET Hospital is providing CBHI based health service since 2019 and the number of beneficiaries of the scheme per month has reached 1500 AaBET insurance registry,Dec 2022). The number of CBHI utilizers is lower at AaBET partly due to availability of third party insurance system for the clients which serves victims of road traffic accidents.The study was conducted in the time period of June 1, 2023-, July 2023

### 2.2. Study Design

Cross sectional, mixed (quantitative and qualitative), method was implemented simultaneously.

#### 2.2.1. Quantitative section

Cross sectional study was conducted using review of patient charts and registries.

#### 2.2.2. Qualitative section

phenomenological study design was conducted using focused group discussion and indepth interview involving the concerned health care workers, CBHI focal persons and patients’attendants. Interview was conducted on the same patients for whom chart and registry is reviewed to get information in depth

### 2.3. Source population

All CBHI insured patients who have got health care services at SPHMMC and AaBET Hospital.

### 2.4 Study Population

CBHI insured inpatients from SPHMMC and AaBET Hospitals which includes elective admissions for procedures and emergency admissions.

### 2.5 Eigibility/ Inclusion and exclusion criteria

Patients admitted to SPHMMC and AaBET Hospital and CBHI insured were eligibile for the study.

#### 2.5.1 Inclusion Criteria

All patients who were admitted to SPHMMC and CBHI insured were included in the study. Experience in CBHI related tasks (> 6months) for the health care workers

#### 2.5.2 Exclusion Criteria

Patients who were unable to speak due to severity of illness in case adequate information can not be obtained

Patients who were new to CBHI scheme (registered <3 months)

Patients who didn’t utilize CBHI scheme at least once previously

### 2.6 Sample size and sampling techniques

The sample size for quantitative section was determined based on the objectives and the highest of the minimum sample size calculated was used for the study.

#### 2.6.1 Sample Size Determination for the first objective (level of good quality CBHI Utilization)

Estimated population proportion was taken from prvious study undertaken on dropping out of Ethiopia’s community-based health insurance scheme reported that good quality of care on offer at health facilities was 38%(5). Factor sampling was also implemented using an online open epi software application which is validated for two population proportion sample size estimation (https://www.openepi.com/SampleSize/SSCohort.htm. A 95% confidence interval was used with 5% margin of error tolerated,variables having p-value ≤0.05 was declared as significant for the association between independent and out come variables.

Finally, the highest among the calculated samples was 362. By adding 10% possible non response rate, the final sample size was 398.

Sample Size for Qualitative study; Purposive sampling was used to create eight groups of 6 to 7 members for focused group discussion. One indepth interview was conducted on selected key informants per each focused group discussion(total of eight). The key informants were selected purposively based on their higher experience on CBHI scheme based services.

#### 2.6.3 Sampling Procedures

##### Quantitative

Systematic random sampling was implemented using bed number of CBHI insured inpatients. Since the average monthly admitted insured patients were 1250 at SPHMMC (1100) and AaBET(150), the interval (kth) value is obtained by dividing 1250 to the sample size 398 which is 3. Hence, every 3rd patient was requested to give consent for participation in the study.

##### Qualitative

Focused group discussion and indepth interview was conducted by purpsively recruiting the concerned health care workers (≥ 6 month experience on CBHI related services), CBHI focals and patients’attendants. This was guided by semistructured quaestionnaire using open ended questions to identify challenges to good quality of utilization of CBHI by clients. Focused Group Discussion was terminated at saturation point.

#### 2.6.4 Study variables

##### Dependent variables

➢ Perceived Quality of Utilization of CBHI based service by clients (details discussed in operational definition)

##### Independent Variables

➢ Client sociodemographic characterstics : Age,Sex,level of education, income residential area(urban, rural)
➢ Client’s individual factors related to quality of CBHI utilization: distance from the hospitals,time to reach hospitals, knowledge on CBHI, knowlege on benefit package,presence of self or family members comorbid illness, perceiption on CBHI service availability and capacity, Catastrophic expenditure, perceived health status,time to recent visit of hospital.prception about care process,trust in scheme managers,non medical cost,satisfaction with the scheme and client’s spending on non-medical costs
➢ Hospital system and delivery of health care related factors: waiting time to get chart, waiting time to be seen by doctors, waiting time between services(consultation to treatment), facility environment cleanliness, inhospital length of stay,Actual CBHI service availability, leadership, presence of quality standards,availability of labs, capacity of labs, availability of drugs, quality of drugs,extra cost by patient due to unavailable services, reimbursement efficiency for extra payment which should have been covered by CBHI, information availability on services,ease of admission(getting beds) and ease of discharge process
➢ Health care workers related factors: perception of interpersonal relations of patients with providers, perception of favor among insured and uninsured, patient volume to providers number, respectful care, timely reponsiveness, perceived experience of health care workers.

### 2.7. Operational definitions of variables

#### Quality of health service utilization

Is delivery of health care at SPHMMC and AaBET hospital to the standard.The World Health Organization (WHO) defines quality of health care as “the extent to which health care services provided to individuals and patient populations improve desired health outcomes” (11)

#### Perceived Good quality ofCBHI based service utilization

those patients who scored above or equal to the mean for standard CBHIS service delivery and related quality indicator questions (Quantitative section,Question number 47-51 which used likert scale to assess, overall timely nature of services,quality of pharmacy, quality of laboratory, quality of administrative services, compassionate and respectfulness and client satisfaction)

#### Perceived Poor quality CBHI based service utilization

those patients who scored below the mean for standard CBHI service delivery and related quality indicator questions.

#### Standard CBHI based health service

This is an essential basic health service provided with out paying out of pocket for CBHI insured patients with premium of 500ETB (10USD) per year for one household. All health services except those are mentioned not to be covered by national CBHIS guideline (e.g CKD dialysis, AKI dialysis >3 months), are expected to be covered by CBHI scheme (admission fee for bed, therapeutic and diagnostic) free of charge keeping the quality standards.

Clients: Insured Patients who were admitted and their guardians who and attend them at SPHMMC or AaBET Hospital during the study period.

#### Barriers to quality of CBHI

Factors identified (in literature) or identified (from this study) to have a negative effect or negative association to quality of utilization by clients.

### 2.8 Data collection Procedures and tools

The Quantitative data was collected from clients at St Paul’s hospital millennium medical college and AaBET hospitals inpatient departments using structured interview. The questionnaire was prepared from standardized MOH and WHO’s CBHI scheme quality indicators as well as peer reviewed journal articles in the literature reviews. In addition, based on the objective of the research relevant questionnaire components were added (Questionnaire is adapted)(10,12,13). The questionnaire was initially prepared in English and translated to Amharic and Afaan Oromo in soft copy. Three smart phones were used for online data collection using Open Data Kit (ODK) soft ware. The data collectors were selecting the type of language as needed and the data was filled by the three trained data collectors. One supervisor and the principal investigator were monitoring the process for completeness, consistency and any challenges and took corrective measures accordingly. The enrolled clients were interviewed and their respective charts and registries were reviewed at least 1 week after admission or on discharge(exit) which ever comes first.This was to avoid respondent bias due to fear of patients to provide genuine information while on care. The qualitative data was collected using audio recorder for focused group discussions and key informant interviews which was transcribed and translated.

### 2.9. Data Analysis Procedure

Separate analysis was conducted for quantitative and qualitative sections.

For the quantitative section :the collected data with ODK was verified for completeness and then it was exported to Excel, coded and then again exported to SPSS Version 25.

Categorization of continuous variables was undertaken before initiation of analysis. A descriptive statistics and frequencies were used for descriptive results. Both binary logistic regression analysis (COR) and multivariable logistic regression (AOR) were performed to assess the association between outcome(dependent)and independent variables. Variables having p value <0.05 were declared as significantly associated with outcome variables and 95% Confidence interval (CI) was used with the odds ratios.

For the qualitative section: Data was first recorded,transcribed, coded and categorized in themes, NVIVO soft ware version 12 was used for analysis in themes. This was conducted using mixed deductive and inductive analysis methods.Then the major reflections and key points of discussants and key informants reported in themes.

Finally, the results were presented by using tables, graphs and texts for the quantitative and using text for the qualitative.

### 2.10 Data Quality Management

The questionnaire was pretested and validated on 40(10 %) of the sample size in St. Petros Hospital, a similar public hospital in Addis Ababa on similar patient groups (insured and admitted inpatients) before starting actual data collection on for the study. Questionnaire was modified based on the result of pretest and the validity test result of the questionnaire was acceptable. We measured the variables on a 5 Likert scale using a 5-point response from very poor to very good, and the internal consistency of the items was checked using the pilot study (Cronbach’s alpha ðαÞ =0:86). The data collectors and supervisor were trained on objectives, procedures, ethical stand of the study and operation of ODK tool for two days For Quantitative section,ODK tool was used with restrictions like escape options,non editability after submission and data collected daily was centrally monitored. The supervisor and principal investigator were actively checking on data for clarity, completeness and consistency. Patient interview was conducted on exit or at least after 1 week of admission. Back up memory was used to retrieve data in case of data loss and the collected data was up loaded online keeping confidentialty and locked with password to minimize data loss. Individual-level data were collected through faceto-face interviews. The data collectors submitted the completed forms to a principal investigator every day, which allowed us to review the submissions and streamline the supervision process.

## 3. Results

5.1 Quantitative section

5.1.1 Socio-demographic characterstics of the study participants.

From a total of 396 participants, 209 (52.8%) were males and 187 (47.2%) were females. In this study group, the mean age was 43.64 years, with a standard deviation of 14.334 years. Most of the participants were urban residents 281(71%), and more than half (53%) were married. More over, most of the respondents (79.3%) were educated to the level of high school or college, whereas the median monthly income was 5000 ETB ranging from 1000 to 15000 ETB. (see the table 1 bellow)

**Table 1:**
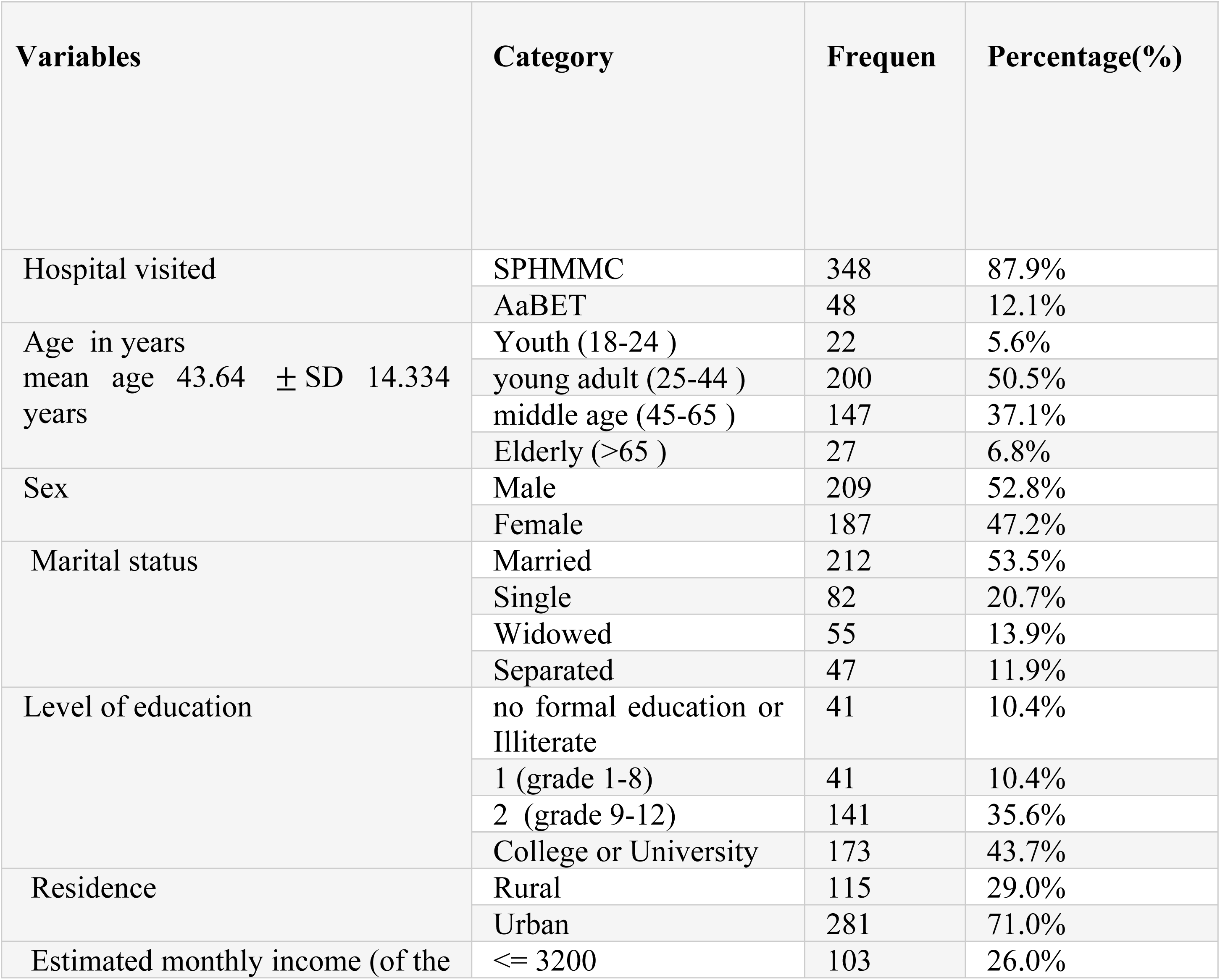
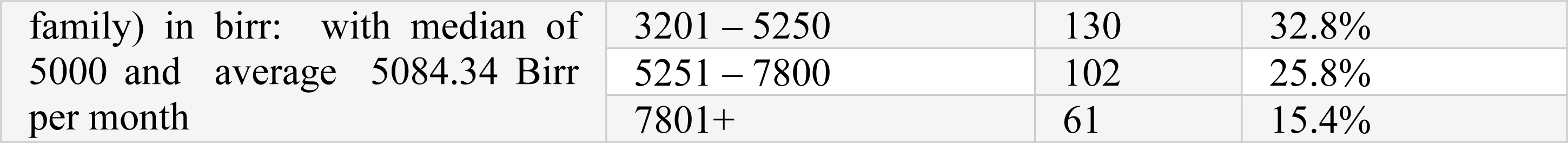
Socio-demographic characterstics of CBHI inpatient utilizers at SPHMMC and AaBET Hospitals, Addis Ababa, Ethiopia from June2023-July 2023.

### 3.1.2. Participant client’s individual factors related to quality of CBHI utilization

In this study, 251 (63%) of participants mentioned that they had faced distance-related difficulties for visiting St.Paul’s Hospital, Millenium Medical College, or AaBET Hospital. The initial enrollment process at community level after subscription to the CBHI scheme was rated as difficult in 38.1% and very difficult in 5.3% of the study participants. Close to two-thirds (287, 72.5%) were satisfied and 32, 8.1%) were very satisfied with the initial enrollment process to the scheme. About 256 (64.6%) of the participants reported that they have faced catastrophic health expenditures in the last two years. After admission to St.Paul’s Hospital, Millenium Medical College, or AaBET Hospitals, 121 (30.6%) of respondents reported that the non-medical expenditure of attendants and patients was costy as detailed in Table 2 bellow

**Table 2:** Description of participant client’s individual factors related to quality of CBHI utilization at SPHMMC and AaBET Hospitals, Addis Ababa, Ethiopia from June 2023-July 2023.

| Variables | Category | Frequency (N) | Percentage (%) |
| --- | --- | --- | --- |
| Challenges related to distance from the hospital | YES | 251 | 63.4% |
|  | NO | 145 | 36.6% |
| Ease of enrolement by the subscriber at Kebele/Woreda: | Difficult/Very difficult | 172 | 43.4 |
|  | Neutral | 46 | 11.6 |
|  | Easy/Very easy | 178 | 44.9 |
| Presence of chronic illness(self or family) | Abscent | 299 | 75.5% |
|  | Present | 97 | 24.5% |
| Previous Frequency of episodes of illness | Moderate (once in 6 months) | 166 | 41.9% |
|  | Frequent (twice in 6 months) | 15 | 3.8% |
|  | Rare (once or none in 12 months) | 215 | 54.3% |
| Experience of catastrophic health expenditure by the family in the last 2 years | No | 140 | 35.4% |
|  | Yes | 256 | 64.6% |
| Satisfaction towards CBHI scheme during enrollment at community level: | Very Dissatisfied | 3 | 0.8% |
|  | Dissatisfied | 32 | 8.1% |
|  | Neutral | 42 | 10.6% |
|  | Satisfied | 287 | 72.5% |
|  | Very satisfied | 32 | 8.1% |
| Non medical expenditure while on admission in the Hospitals: | Affordable | 275 | 69.4% |
|  | Costly | 121 | 30.6% |

This study has shown that the knowledge status among Admitted Patients towards community-based health insurance was revealed to be 144 (36.40%) having good knowledge, followed by 233 (58.80%) having moderate knowledge towards community-based health insurance, In this study 251(63%) participants complained that delay in getting chart and 235(59%) stated delay to be seen by doctors. About 145(36.4%) particpants ware gating chart timely with out delay.

In addition, 336 (84.4%) participants mentioned that drug availability was inadequate and suffering from stock outs at St.Paul’s Hospital, Millenium Medical College, or AaBET hospitals. On the other hand, about 270 (67.8%) participantes rated the available drugs as agood quality interms of efficacy and availability of first line drugs as described by their physicians where as the rest 32.2% rated as poor.

This study also revealed that 277 (69.6%) of participants had faced shortage of laboratory services at the hospitals. About 209(52.5%) of participants faced challenges form unavailable radiology services after admission to the hospitals.

Turn around time is the time needed for result acquisition after laboratory or radiology service is ordred and the process is initiated. In this study, 44.4%(177) and 198(49.7%) of the participants reported that the lab and radiology results were delayed respectively.

Appart from that, most of respondants complained that medications and services which were unavailable in the hospital but available from private were too expensive. 44% of the respondents mentioned challenges related to admission to inpatient (bed availability and process).

### 3.1.3. Over all quality of CBHI scheme based services in the hospitals

The over allquality of CBHI scheme based health service in the study hospitals was determined using the mean of six quality indicator variables (Q46-51) including over all quality of timely nature of services, laboratory services, pharmacy services, administrative services,compassionate and respectful services) and satisfaction with hospital services.

This study determined the overall Quality of community-based health insurance utilization among admitted patients at SPHMMC and AaBET hospitals revealing that 288 (72.7%) of admitted patients utilized good Quality community-based health insurance services, with 95% CI (68 to 77).

#### 3.2. Factors associated with good quality of CBHI utilization

Model fitness was tested with Hosmer-Lemeshow goodness of fit test (0. 881). Logistic regression model were used to compute Bivariable and multivariable analysis. The Bivariable analysis was used to measure the association between each independent and dependent variables. Those variables that have P-value < 0.25 on bivariate analysis were entered into a multivariable logistic regression model to control for confounder and identify the independent predictors of good quality of CBHI utilization. Bivariable logistic regression analysis identified factors significantly associated with quality of CBHI utilization at a p-value < 0.25 were : facility(Hospital) environment is clean and attractive,waiting time to get chart in CBHI on arrival to this hospital, length of stay in hospital in days, availability and access of information on CBHI service, adequate number of health professionals who provide services for patient volume, challenges due to distance from this hospital, family income,knowledge towards CBHI scheme, ease of enrollment (premium fee and process) at subscription for CBHI, and overall satisfaction towards CBHI at community level during enrollment. (See the table 3 bellow)

**Table 3:**
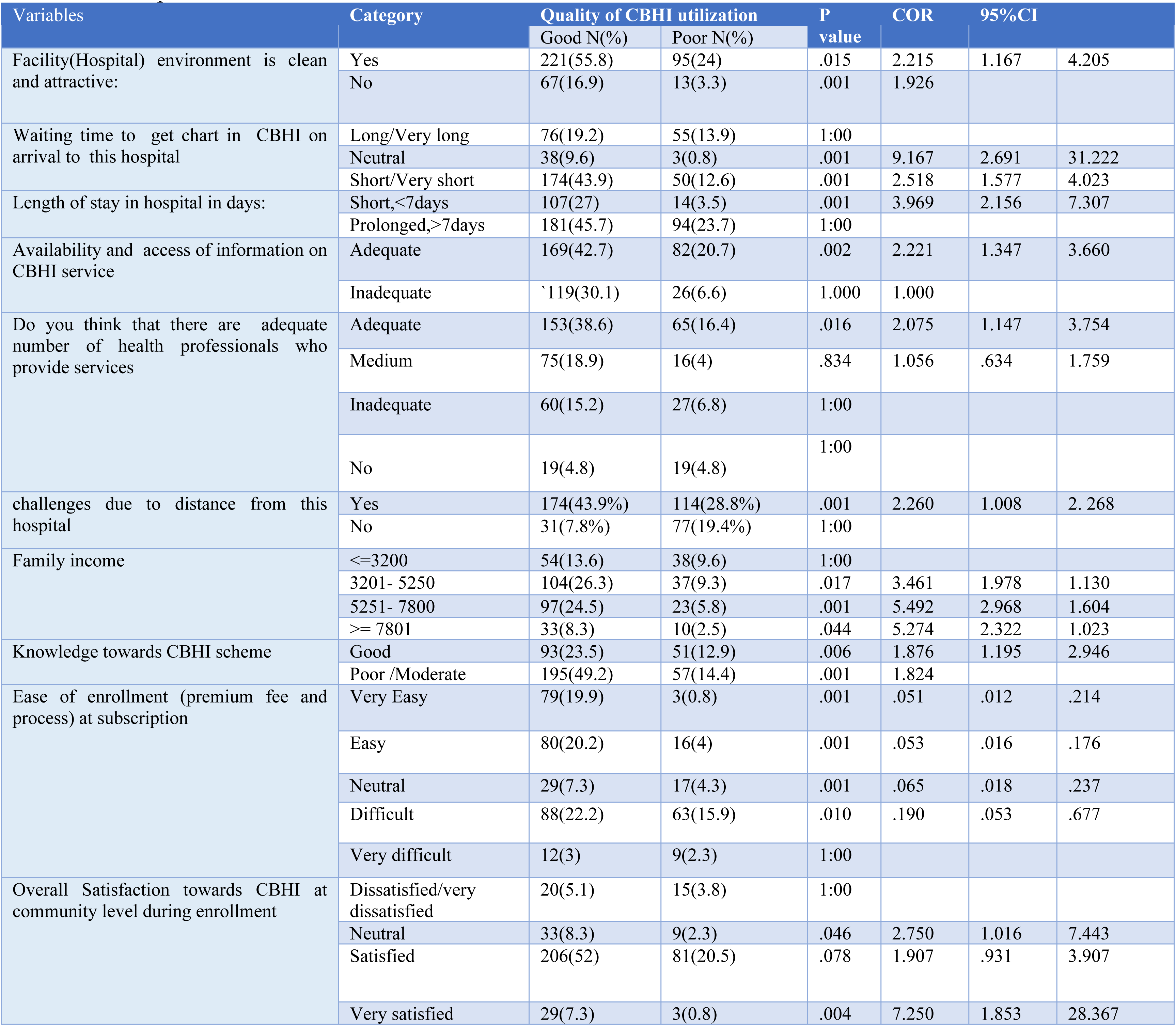
Identified factors significantly associated with Quality of CBHI utilization at Bivariable logistic regression analysis among participants at SPHMMC and AaBET Hospials Addis Ababa, Ethiopia 2023.

| Variables | Category | Quality of CBHI utilization |  | P value | COR | 95%CI |  |
| --- | --- | --- | --- | --- | --- | --- | --- |
|  |  | Good N(%) | Poor N(%) |  |  |  |  |
| Facility(Hospital) environment is clean and attractive: | Yes | 221(55.8) | 95(24) | .015 | 2.215 | 1.167 | 4.205 |
|  | No | 67(16.9) | 13(3.3) | .001 | 1.926 |  |  |
| Waiting time to get chart in CBHI on arrival to this hospital | Long/Very long | 76(19.2) | 55(13.9) | 1:00 |  |  |  |
|  | Neutral | 38(9.6) | 3(0.8) | .001 | 9.167 | 2.691 | 31.222 |
|  | Short/Very short | 174(43.9) | 50(12.6) | .001 | 2.518 | 1.577 | 4.023 |
| Length of stay in hospital in days: | Short,<7days | 107(27) | 14(3.5) | .001 | 3.969 | 2.156 | 7.307 |
|  | Prolonged,>7days | 181(45.7) | 94(23.7) | 1:00 |  |  |  |
| Availability and access of information on CBHI service | Adequate | 169(42.7) | 82(20.7) | .002 | 2.221 | 1.347 | 3.660 |
|  | Inadequate | 119(30.1) | 26(6.6) | 1.000 | 1.000 |  |  |
| Do you think that there are adequate number of health professionals who provide services | Adequate | 153(38.6) | 65(16.4) | .016 | 2.075 | 1.147 | 3.754 |
|  | Medium | 75(18.9) | 16(4) | .834 | 1.056 | .634 | 1.759 |
|  | Inadequate | 60(15.2) | 27(6.8) | 1:00 |  |  |  |
|  | No | 19(4.8) | 19(4.8) | 1:00 |  |  |  |
| challenges due to distance from this hospital | Yes | 174(43.9%) | 114(28.8%) | .001 | 2.260 | 1.008 | 2. 268 |
|  | No | 31(7.8%) | 77(19.4%) | 1:00 |  |  |  |
| Family income | <=3200 | 54(13.6) | 38(9.6) | 1:00 |  |  |  |
|  | 3201- 5250 | 104(26.3) | 37(9.3) | .017 | 3.461 | 1.978 | 1.130 |
|  | 5251- 7800 | 97(24.5) | 23(5.8) | .001 | 5.492 | 2.968 | 1.604 |
|  | >= 7801 | 33(8.3) | 10(2.5) | .044 | 5.274 | 2.322 | 1.023 |
| Knowledge towards CBHI scheme | Good | 93(23.5) | 51(12.9) | .006 | 1.876 | 1.195 | 2.946 |
|  | Poor /Moderate | 195(49.2) | 57(14.4) | .001 | 1.824 |  |  |
| Ease of enrollment (premium fee and process) at subscription | Very Easy | 79(19.9) | 3(0.8) | .001 | .051 | .012 | .214 |
|  | Easy | 80(20.2) | 16(4) | .001 | .053 | .016 | .176 |
|  | Neutral | 29(7.3) | 17(4.3) | .001 | .065 | .018 | .237 |
|  | Difficult | 88(22.2) | 63(15.9) | .010 | .190 | .053 | .677 |
|  | Very difficult | 12(3) | 9(2.3) | 1:00 |  |  |  |
| Overall Satisfaction towards CBHI at community level during enrollment | Dissatisfied/very dissatisfied | 20(5.1) | 15(3.8) | 1:00 |  |  |  |
|  | Neutral | 33(8.3) | 9(2.3) | .046 | 2.750 | 1.016 | 7.443 |
|  | Satisfied | 206(52) | 81(20.5) | .078 | 1.907 | .931 | 3.907 |
|  | Very satisfied | 29(7.3) | 3(0.8) | .004 | 7.250 | 1.853 | 28.367 |

The multivariable logistic regression analysis identified that factors significantly (P<0.05) with the quality of CBHI utilization were: facility (hospital) environment is clean and attractive; satisfaction towards CBHI at community level, knowledge towards the CBHI scheme; availability and access of information on CBHI service, family monthly income as well as welcoming, respectful, and compassionate CBHI service is provided.

Community-based health insured inpatients who responded that the hospital environment was clean and attractive had 2.8 times higher odds of utilizing good-quality CBHI service as compared to poor-quality CBHI service utilizers [AOR = 2.77:95% CI (1.24–6.165)].

Community-based health insurance members who were satisfied during enrollment at community level among the admitted patients were 2.45 times [AOR = 2.45:95% CI (1.11–5.39)], and very satisfied patients were 8.8 times [AOR = 8.79:95% CI (2.12–36.44)] more likely to utilize good-quality CBHI services as compared to dissatisfied or very dissatisfied community-based health insurance members.

Community-based health insurance members of admitted patients who had good knowledge of the CBHI scheme were twice as likely [AOR = 1.97:95% CI (1.2–3.23)] to utilize good-quality CBHI services as compared to their counterparts.

The odds of adequate availability and accessibility of information on CBHI services were increased by 2.4 times for good-quality CBHI services compared to inadequate CBHI information [AOR = 2.37:95% CI (1.34–4.21)].Community-based health insured members with family monthly income between 5251-7800 and greater than 7801 birr were 2.6 times [AOR = 1.97:95% CI (1.2–3.23)] and 5.3 times [AOR = 5.3:95% CI (2.32–10.23)] more likely to use good-quality CBHI services compared to those with less than 3200 birr per month. (see the table 4 bellow)

**Table 4:** Bivariable and multivariable logistic regression analysis identified factors significantly associated with Quality of CBHI utilization among admitted patients at SPHMMC and AaBET Hospials Addis Ababa, Ethiopia 2023 (n = 396)

| Variables | Category | Quality of CBHI utilization |  | P value | AOR | 95%CI |  |
| --- | --- | --- | --- | --- | --- | --- | --- |
|  |  | Good N(%) | Poor N(%) |  |  |  |  |
| Facility(Hospital) environment is clean and attractive: | Yes | 221(55.8) | 95(24) | .013 | 2.767 | 1.242 | 6.165* |
|  | No | 67(16.9) | 13(3.3) | 1:00 |  |  |  |
| Satisfaction towards CBHI at community level | Neutral | 33(8.3) | 9(2.3) | .231 | 1.958 | .651 | 5.886 |
|  | Satisfied | 206(52) | 81(20.5) | .027 | 2.447 | 1.110 | 5.396* |
|  | Very satisfied | 29(7.3) | 3(0.8) | .003 | 8.792 | 2.122 | 36.435** |
|  | Dissatisfied/very dissatisfied | 20(5.1) | 15(3.8) | 1:00 |  |  |  |
| Knowledge towards CBHI scheme | Good | 93(23.5) | 51(12.9) | .007 | 1.970 | 1.200 | 3.233** |
|  | Poor /Moderate | 195(49.2) | 57(14.4) | 1:00 |  |  |  |
| Availability and access of information on CBHI service in the hospitals | Adequate | 169(42.7) | 82(20.7) | .003 | 2.374 | 1.339 | 4.210** |
|  | Inadequate | 119(30.1) | 26(6.6) | 1:00 |  |  |  |
| Family income | <=3200 | 54(13.6) | 38(9.6) | 1:00 |  |  |  |
|  | 3201- 5250 | 104(26.3) | 37(9.3) | .155 | 1.555 | .847 | 2.855 |
|  | 5251- 7800 | 97(24.5) | 23(5.8) | .004 | 2.623 | 1.350 | 5.095** |
|  | >= 7801 | 33(8.3) | 10(2.5) | .044 | 5.274 | 2.322 | 1.023* |

Note: - \*\*\*shows significant at P-value < 0.001 , \*\*P-value < 0.01 & \*shows significant at P-value < 0.05, Abbreviation: CBHI - community-based health insurance, , COR: Crude Odd Ratio, AOR: Adjusted Odd Ratio, CI: Confidence Interval. 1:00 means reference category

## 4. Qualitative analysis results

### Socio demographic characterstics of participants

A total of 8 focus group discussions were conducted with 6 to 7 members per group. The groups characterstics are depicted in table 5 below

**Table 5:** Socio demographic characterstics of participants on qualitative study.

| FGD No | FGD participant list | Mambers of each group | Age group | Sex |
| --- | --- | --- | --- | --- |
| 1 | SPHHMC CBHI | 6 | 27-46 | 2 of them are females |
| 2 | LIASION (SPHHMC) | 6 | 26-50 | 3 of them are males |
| 3 | Icu and supervision (SPHHMC) | 6 | 28-40 | 1 of them are female |
| 4 | Health workers SPHHMC | 7 | 25-32 | 4 of them are males |
| 5 | Attendants (SPHHMC) | 7 | 18-55 | 2 of them are females |
| 6 | CBHI(AaBET ) | 6 | 32-38 | 2 of them are males |
| 7 | LIASION(AaBET) | 6 | 29-43 | 3 of thm are males |
| 8 | Attendants(AaBET) | 6 | 23-57 | 3 of them are males |

### 4.1. Qualitative thematic analysis

#### ThemeI: Barriers to Quality of CBHI utilization related to hospital infrastructure and supply chain availability

Participant 1 from group1(P1G1) from CBHI group said that “Many clients face challenges related to hospital structure and failure to provide clear guidance with signet to their service areas which is also true at St.Pauls.” This was also noted by P4 from liaison group who stated “Patients often face difficulties in searching through the hospital system and service locations leading to a inconvienence in accessing the healthcare services.”

In addition, regarding the challenges of drug shortages and frequent stock outs in the hospitals he stated that “Drug shortages and frequent stock outs pose a significant challenge to CBHI utilization leading to delayed or inadequate healthcare. This problem requires attention and solutions to ensure a consistent supply of medications.”

On the other hand, P2 gives a basic information about lack of medications at Kenema pharmacy “while Kenema pharmacy is supposed to avail drugs to CBHI consumers, the lack of medications at Kenema pharmacy is a major problem. Addressing this issue requires improving the supply chain and ensuring a consistent stock of medications at Kenema pharmacy.”

From CBHI groups participant 3 was concerned about ordering and introduction of high-quality drugs to CBHI list “The challenge related to ordering and introducing high-quality drugs to the CBHI list can limit access to essential medications since these drugs are not available in CBHI scheme currently”

Shortage of reagents and laboratory services were also identified by different participants in different groups as “The shortage of reagents and laboratory services are hindering accurate diagnoses and appropriate treatment compromising the quality of care and patient outcomes.”

P4 from CBHI group explains in addition, there is shortage of healthcare professionals compared to patient flow, “The shortage of healthcare professionals, relative to the patient flow, is a major obstacle in maximizing CBHI utilization. This issue highlights the need for increased recruitment and retention of healthcare professionals to meet the growing demand.”

Another participant from different groups said “The increasing patient flow puts strains on healthcare facilities and resources, potentially compromising the quality of care provided. With limited capacity to handle higher patient volumes, healthcare providers might be overwhelmed to meet the needs of **CBHI beneficiaries, leading to longer waiting times and reduced attention to individual patients.hhmm….** Empowering CBHI employees can contribute to improved patient experiences and outcomes.**”**

Most of the participants recommended that addressing these coverage limitations and access difficulties are crucial for ensuring equitable access to healthcare services. Key informant 2“There is a need for continuous adjustments and improvements in health insurance systems, including policy reforms, increased public awareness campaigns, enhanced coordination between insurance providers and healthcare facilities, and ongoing evaluation of the effectiveness of health insurance schemes.” On the other hand,CBHI scheme coverage limitations were reported as barriers to utilization of the scheme. P1 from ICU group and P1 from supervision group of SPHMMC stated “CBHI does not provide coverage for expensive services such as chronic kidney disease (CKD)dialysis,Arterial blood gas, transplants, and some of dental services, which can create financial barriers for patients in need of these services”

#### ThemeII: Challenges related to knowledge of providers on CBHI scheme and communication

Participant six from CBHI team described about lack of understanding of CBHI guidelines and policies, explains as “The lack of understanding of CBHI guidelines and policies at the Woreda (district) level can hinder effective implementation and utilization of CBHI services at different stages including the hospitals. Inconsistent interpretation and application of guidelines may result in variations in care quality and coverage, impacting the overall satisfaction and trust of CBHI beneficiaries.” This is related to misunderstanding among health care providers as well. For instance, a participant from ICU team reported that “imaging services including x-ray are not covered under CBHI scheme” which was contradicting to the CBHI guideline which allows free coverage of these imaging services under CBHI scheme.

From CBHI groups participant 3 said that lack of communication with users regarding changes in guidelines and policies is abarrier on quality of services: “Failure to effectively communicate changes in CBHI guidelines and policies to users can lead to confusion and misunderstandings. This lack of communication may result in beneficiaries not receiving the appropriate care or being unaware of important updates, potentially compromising the quality of their healthcare experience.”

P3 from AaBET hospital CBHI group faced problems related to lack of updated and accessible information for users. This was also supported by Key informant 1 from AaBET hospital who recommended improvement in communication stating “Creating effective communication channels and implementing patient-centered services can facilitate better engagement, understanding, and satisfaction among CBHI beneficiaries”.

#### Theme III Problems with individual Community-Based Health Insurance (CBHI) users

CBHI users often face various challenges related individual personal characterstics P1 from attendants faced lack of knowledge and awareness about CBHI coverage: “Many individuals are unaware of the extent of CBHI coverage and the specific services and benefits they are entitled to, leading to underutilization of the program and missed opportunities for accessing necessary healthcare”

key informant 3 from ICU described the impact of knowledge gap as “Many users of Community-Based Health Insurance (CBHI) lack the necessary training and knowledge about the coverage provided by the insurance scheme, leading to confusion and potential difficulties in accessing appropriate healthcare services.”

Similarly a key informant from CBHI group of SPHMMC has raised knowledge gap as a challenge along with other factors “By addressing difficulties faced by users, increasing knowledge and awareness, streamlining reimbursement processes, improving access to services and addressing resource limitations, CBHI can better serve the needs of the community and improve healthcare outcomes. The quality of healthcare services covered by insurance plans can vary, and patients may face difficulties related to long waiting times, inadequate medical facilities, or unsatisfactory treatment outcomes rsulting in poor perception of the service quality.”

“The absence of updated and easily accessible information about hospital services, appointment schedules, and medical procedures adds to the inconvenience faced by patients. It is essential to provide patients with adequate information regarding their healthcare options, treatment plans, and any relevant guidelines to ensure informed decision-making and active participation in their care.”Participants also suggested addressing communication challenges, such as language barriers, unclear instructions, or poor doctor-patient communication, is essential for effective healthcare delivery and patient understanding.

From SPHMMC supervision office P4 faced problems related to distance of the tertiary hospitals from patients:”The inconvenient location of some hospitals makes it challenging for individuals residing far from the city center to access necessary medical facilities.”

P3 from ICU faced problems of inconvenience in accessing services outside of Addis Ababa as “The limited availability of healthcare services outside of Addis Ababa creates difficulty for individuals living in rural areas who have to travel long distances to receive specialized medical care.”

#### Theme IV :Administrative challenges in CBHI utilization

P1 from attendants faced difficulty during stamping and activation of CBHI cards “The process of stamping and activating CBHI cards can present challenges, potentially leading to delays or complications in accessing healthcare services”. P 2 from patient attendant was also complaining on the same issue stating “Patients experience significant waiting times for card issuance and to be seen by health professionals, which can lead to delays in receiving care and potentially affect patient outcomes.”

P2 from CBHI reported overcrowding with insufficient windows and rooms for excess patient flow: “The limited availability of windows and rooms to accommodate the increasing patient flow can create administrative challenges, leading to longer waiting times and potential disruptions in service delivery”. Similarly, CBHI groups P3 experienced long queues and waiting times which was stated as “The overcrowded hospitals result in long queues and waiting times, causing inconvenience for patients who require immediate medical attention which is happening at St.Paul’s”

Addressing these administrative challenges is essential for improving the efficiency and effectiveness of CBHI utilization. This may involve streamlining the stamping and activation process for CBHI cards and ensuring adequate infrastructure to handle the growing patient flow. P2 from CBHI recommended that “The implementation of Electronic Medical Records (EMR) can enhance efficiency, accuracy, and continuity of care within the CBHI system”

#### Theme V challenges with hospital services and bed availability

P4 from attendants was complaining about the difficulty in finding beds and accessing certain services:“There are challenges in finding beds and accessing specific services within the hospital, which can result in delayed or limited access to appropriate care”.

P1 from patient attendants “complains about cold and discomfort while waiting in the emergency room”. Patients have expressed complaints regarding the cold and discomfort they experience while waiting in the emergency room for inpatient admission which created discomfort and decreased overall satisfaction with the healthcare facility..

Key informant 3 from ICU team “To address these barriers, several remedial measures are suggested. These include recognizing and involving the community in decision-making processes, empowering healthcare professionals through training and support, reducing financial barriers by making healthcare services more affordable, focusing on recruiting and retaining skilled professionals to improve service delivery, and reminding the institution to promptly address problems on time”.

P2 from patient attendants group also faced problems with getting money back from CBHI after payment at private. He “almost all CBHI users encountered difficulties in the reimbursement process, experiencing delays in receiving their money back for healthcare expenses covered by the insurance, which created financial burdens and dissatisfaction”.Reimbursement was reported to be implemented only by some regions of Ethiopia creating inconsistency in CBHI scheme approach.

Addressing these challenges is crucial for improving the quality of hospital services. Strategies proposed by FGD members and key informants include; streamlining administrative processes to reduce waiting times, optimizing bed management systems, and improving the comfort and amenities provided to patients in waiting areas.The overall information suggests that addressing the gaps in CBHI service provision, user and provider training, improving communication and information dissemination are needed. The findings highlight, the importance of community awareness, ensuring users have a clear understanding of coverage and exceptions, and finding solutions to financial constraints.

#### Strength and Limitation of the study

This study is one of few studies conducted at specialized or tertiary hospital levels applying both quantitative and qualitive mixed design implementaing method. The sudy has identified important barriers to good quality CBHI utilization which are poteintially changeable.

How ever, the study is cross sectional that it may not show cause and effect relation between dependent and independent variables. This study was also conducted in limited facilities SPHMMC and AaBET indicating the need for further multicenter researches.

#### Conclusion and recommendation

This study revealed that more than two third of admitted patients at SPHMMC and AaBET hospital utilized good qulity community-based health insurance(CBHI) services which is slightly higher than most of previous studies.

Factors significantly affecting quality CBHI utilization included a clean and attractive facility environment, satisfaction towards the CBHI scheme at community level, knowledge of the scheme, availability and access to information as well as family monthly income. These factors contribute to the overall quality of CBHI utilization. This study has also identified presence of major challenges related to shortage of drugs and cost of unavailable drugs and lab services to the CBHI consumers to purchase from private facilities coupled with lack of reimbursement.

Qualitatively, this study explores the barriers to quality of Community Based Health Insurance utilization, in relation to facility environment, infrastructure administrative system of the scheme, health care workers and individual clients related barriers at SPHMMC or AaBET Hospital. It also seeks suggestions on how to improve CBHI utilization at these hospitals, focusing on facility system, health care providers and client-related factors identified from qualitative findings.

Hence, its recommended that hospital mangers, CBHI facilitators at community level, health care providers, ministry of health and CBHI organization focus on the aforementioned factors to improve the quality of CBHI scheme based service and further multicenter studies are warranted particularly at general or tertiary hospitals.

## Data Availability

we will submit it up on request

## Acronyms and Abbreviations

AaBET: Hospital Addis Ababa Burn, Emergency and Trauma Hospital
AOR: Adjusted Odd’s Ratio
CBHI: Community Based Health Insurance
CBHIS: Community Based health Insurance scheme
CKD: Chronic Kidney Disease
FGD: Focused Group Discussion
HSTP II: Health Sector Transformation plan II
ICU: Intensive care unit
LMIC’S: Low and Middle Income Countries
MOH: Ministry of Health
PQoC: Perceived Quality of Care
SPHMMC: Saint Paul’s Hospital Millennium Medical College
UHC: Universal Health Coverage
SHI: Social health insuranc

## Declarations

### -Ethics approval and consent to participate

Ethical clearance was obtained from the research and ethics review board of St. Paul’s Hospital Millennium Medical College with Ethical Review Board Ref. No. PM23/713. The respective hospitals were communicated and permission was obtained from the respective hospital’s responsible bodies before data collection. Confidentiality of information was maintained throughout the study process. Participation in this study was voluntary and participants had full right not to participate or withdraw from the study after signing on written informed consent. There was no personal relation and no conflict of interest between the authors, supervisors, facilitators, and participants. The potential risk of participation in this study is negligible.

### -Consent for publication

All authors gave their consent for this original article to be published on international journal

### -Availability of data and materials

The soft copy and data for this research is avialble and will be submmited by the corresponding author up on request

### Authors’ contributions

**D.K.** contributed to the conception, design, questionry development, drafting the research proposal and conduct of the study. **Y.L** participated in research tool selection and development, data collection, research analysis, manuscript preparation and submission. **A.G and T.G**. contributed to the design and conduct of the study. All authors analyzed and interpreted the data, read and approved the final manuscript

## Acknowledgements

We would like to extend our sincere gratitude to the study participants, data collectors without whom this study could have been impossible. It is with honor that we would like to appreciate St. Paul’s Hospital Millennium Medical College, School of Public Health for giving us an excellent opportunity to practice research and guiding me starting from development proposal through completion of this study.

## -Authors’ information

Not applicable

## -Availability of Data and Materials

is available and will be disclosed up on request from the corresponding author

## -Consent for publication

Not Applicable for this section

